# Feasibility and Preferences for Home-based Self-Testing for HIV, Diabetes, and Hypertension in Kenya, South Africa, and Zambia

**DOI:** 10.1101/2025.10.28.25339020

**Authors:** Carolyn Oliver, Meagan J. Bemer, Amber Lauff, Jennifer F. Morton, Shawna Cooper, Hilton Humphries, Anjali Sharma, Derek Pollard, Anna Winters, Alastair van Heerden, Elizabeth Bukusi, Dino Rech, Paul K. Drain

## Abstract

Home-based self-testing may improve individual health outcomes and public health disease surveillance by lowering the barriers associated with clinic-based diagnostic testing in low- and middle-income countries (LMICs). We assessed the feasibility of conducting home-based, self-testing for key communicable and non-communicable diseases, including HIV, diabetes, and hypertension, in sub-Saharan Africa. We enrolled participants (≧15 years) from households in peri-urban and rural communities of Kenya, South Africa, and Zambia. Participants opted in to self-directed rapid testing for HIV and blood glucose and had their blood pressure measured by a research team member. Our primary measures included HIV status and testing history, HIV and blood glucose rapid test results, blood pressure, self-reported usability and acceptability of self-testing, and participant preferences for future self-testing. Among the 526 participants from 100 households enrolled in each country, the average age was 41 years and 63% were female. Overall, 16% of participants reported living with HIV. Over half of participants (52%) had last tested for HIV >12 months ago or had never tested for HIV, and 8% of participants were unsure of their HIV status. Among participants who self-tested, 2% (N=6) tested positive for HIV and 4% (N=18) had high blood glucose, while 26% (N=131) had high blood pressure measured by the clinical team. Only 13% of study participants reported previously using a rapid test. Most (>90%) participants rated all procedures for HIV and blood glucose tests as either “very easy” or “fairly easy” to use. Most participants (88%, N=458) preferred home-based testing. Home-based self-testing for HIV and blood glucose and testing for blood pressure were feasible and preferred in peri-urban and rural areas of Kenya, South Africa, and Zambia. Self-testing has potential to expand and accelerate access to healthcare delivery in LMICs.

## Introduction

In low- and middle-income countries (LMICs), the high burden of acute infections [1] (e.g., HIV, TB, malaria, COVID-19) and the growing prevalence of non-communicable diseases (NCDs) [2] (e.g., diabetes, hypertension, heart disease) pose challenges for healthcare delivery. [3,4] Barriers to clinic-based testing include cost of transportation, long wait times, lost results, inadequate clinic staffing, lack of consistently available diagnostic test stock, and perceived stigma. [5–9] At-home self-testing offers a promising alternative to clinic and hospital-based care by increasing privacy and encouraging earlier diagnostic testing, which can reduce time to seek and receive treatment – with particularly encouraging developments in HIV diagnosis and treatment. [10–12] Advances in diagnostics, like rapid tests, some of which are approved for self-testing, can help expand and accelerate access to healthcare delivery, while empowering individuals with self-care.

In 2024, the World Health Organization (WHO) published guidelines [13] to advance self-care programs that lower healthcare costs and increase access to care by prioritizing individual agency and self-determination. Self-testing using rapid tests was proposed as one of several healthcare tools that patients could use effectively for self-care. Self-testing offers population health benefits, including expanding access to healthcare services for women, who often face barriers to clinic-based care. [14,15] Self-testing studies have demonstrated that individuals without medical training can reliably conduct and interpret their own rapid test results [16,17] including in LMICs, albeit primarily demonstrated in single-site studies for a single communicable disease. [10,18–23]

We assessed the feasibility of conducting home-based, self-testing for communicable and NCDs in multiple peri-urban and rural communities of sub-Saharan Africa, and to estimate the prevalence of HIV, high blood glucose, and high blood pressure in our study population.

## Materials and methods

### Study design, sites and partners

The DASH (Diagnostic Access to Self-Care & Health Services) Study was a cross-sectional study evaluating the feasibility of home-based testing via household survey and provision of self-testing tools in Kenya, South Africa, and Zambia. Peri-urban communities with populations of 200 to 2,000 people were chosen within each study site, which included Migori County, Kenya; KwaZulu-Natal Province, South Africa; and Luanshya District, Copperbelt Province, Zambia.

Migori County, Kenya is a peri-urban county of 1.1 million people and faces high burdens of HIV and malaria. Umsunduzi sub-district in KwaZulu-Natal province, South Africa, has approximately 618,000 people living in urban, peri-urban, and rural areas and has a high burden of HIV, sexually transmitted infections (STIs), early pregnancy, and diabetes. Luanshya District, Zambia has 118,000 people and includes peri-urban and rural areas with a high burden of HIV, malaria, and early pregnancy.

The DASH Study used a mobile app, HealthPulse TestNow™ (Audere; Seattle, USA) to guide study participants as they self-tested for HIV and blood glucose. The app was tailored to each study site, accounting for the types and brands of rapid tests used and the local language.

### Methodology

Household surveys were conducted by research team members in each of the study sites. Participants were considered members of a household if they spent at least one night there in the past four weeks.

Participants aged 6 months and greater were eligible to be enrolled in the at all study sites in the DASH Study. This analysis focused on adolescents and adults aged ≥15 years old. Individuals 16 years and older provided informed written consent to participate, while individuals 15 years of age were required to provide parental consent to participate in the study. The research team collected socio-demographic data and current medical conditions on encrypted tablet devices using REDCap Version 14. The study was reviewed by the University of Washington Institutional Review Board, the Human Sciences Research Council (HSRC) Research Ethics Committee in South Africa, the Scientific and Ethics Review Unit at the Kenya Medical Research Institute, and ERES Converge IRB in Zambia.

After conducting the household survey, all participants with or without symptoms were offered a mobile app-facilitated test for HIV and blood glucose. The AI-powered app, HealthPulse TestNow, was available on a study-provided mobile device. The app guided participants through the steps of accurately administering and interpreting each test. The full version of the app included process control timers, the ability to capture a quality photo of the test, result interpretation guidance, and an artificial intelligence algorithm which runs directly on the mobile device and can interpret results from RDT images. For this study, AI interpreted results were not shared with participants. Participants in Kenya had access to the full version of the app, allowing them to take photos of their test result. In Zambia and South Africa, a prototype of the app was used, and participants did not take photos of their test result. Instead, the test result image was recorded by a research team member.

For testing, the Kenya site used Mylan HIV finger prick self-tests, oral OraQuick HIV self-tests, and the On Call Plus glucose testing system. The South Africa site used OraQuick HIV self-tests and the Accu-Chek glucose testing system. The Zambia site used Abbott Determine finger prick HIV tests and the Accu-Chek glucose testing system. Participants at the Zambia site did not collect their own HIV finger prick whole blood specimens due to stock availability of an alternative test approved for self-testing. Instead, the biological specimen was collected and applied to the test for the participant by a research team member. However, participants at the Zambia site were given the study mobile device to follow along in the app and were surveyed on the ease of understanding the instructions.

Participants were asked to interpret the result of the HIV rapid test and report their interpretation to the research team. The research team, who were all trained to accurately interpret the HIV self-tests, also interpreted the test result themselves and recorded the result. After each self-test for HIV and blood glucose levels, participants were asked about the ease of understanding the test instructions and the ease of conducting the test.

Participants had the option to receive a blood pressure measurement by a member of the research team. Digital blood pressure measurement devices were used at each study site, including Omron in Kenya, Contec and Axcess in South Africa, and Citizen in Zambia.

### Sample size and statistical analyses

Sample size at each study site was not powered for statistical significance because the primary measure was feasibility. Each study site had a cutoff of 100 households. In Kenya and Zambia, households were selected by convenience sample and the research teams enrolled as many household members as were able and willing to consent. In South Africa, the research team used the Kish grid method [24] to systematically select a household for participation when more than one household was found at a visiting point, and to randomly select a maximum of three members per household. The process involved creating two separate grids -- one with randomly selected visiting points and the other with the number of members to be selected within a household. The Kish grid method ensures a representative and unbiased sample when conducting surveys or studies across various households.

Descriptive statistics were calculated for all study measures, including percentage, mean, standard deviation, and median with interquartile range using python 3.8.3.

## Results

The research teams conducted 300 household visits, with 100 households at each study site, between 31 May and 4 August 2023, enrolling 526 adolescent and adult participants in Kenya (N=197), South Africa (N=148), and Zambia (N=181) (Table 1). The household composition, including enrolled household members and those who did not participate in the study, was variable across study sites. In Kenya, households had 5 members on average, while South Africa and Zambia had an average of 2 and 3 household members respectively. The Kenya site households had an even number of male and female members (48% female) while South Africa and Zambia had majority female participants at 62% and 65%, respectively.

**Table 1.**
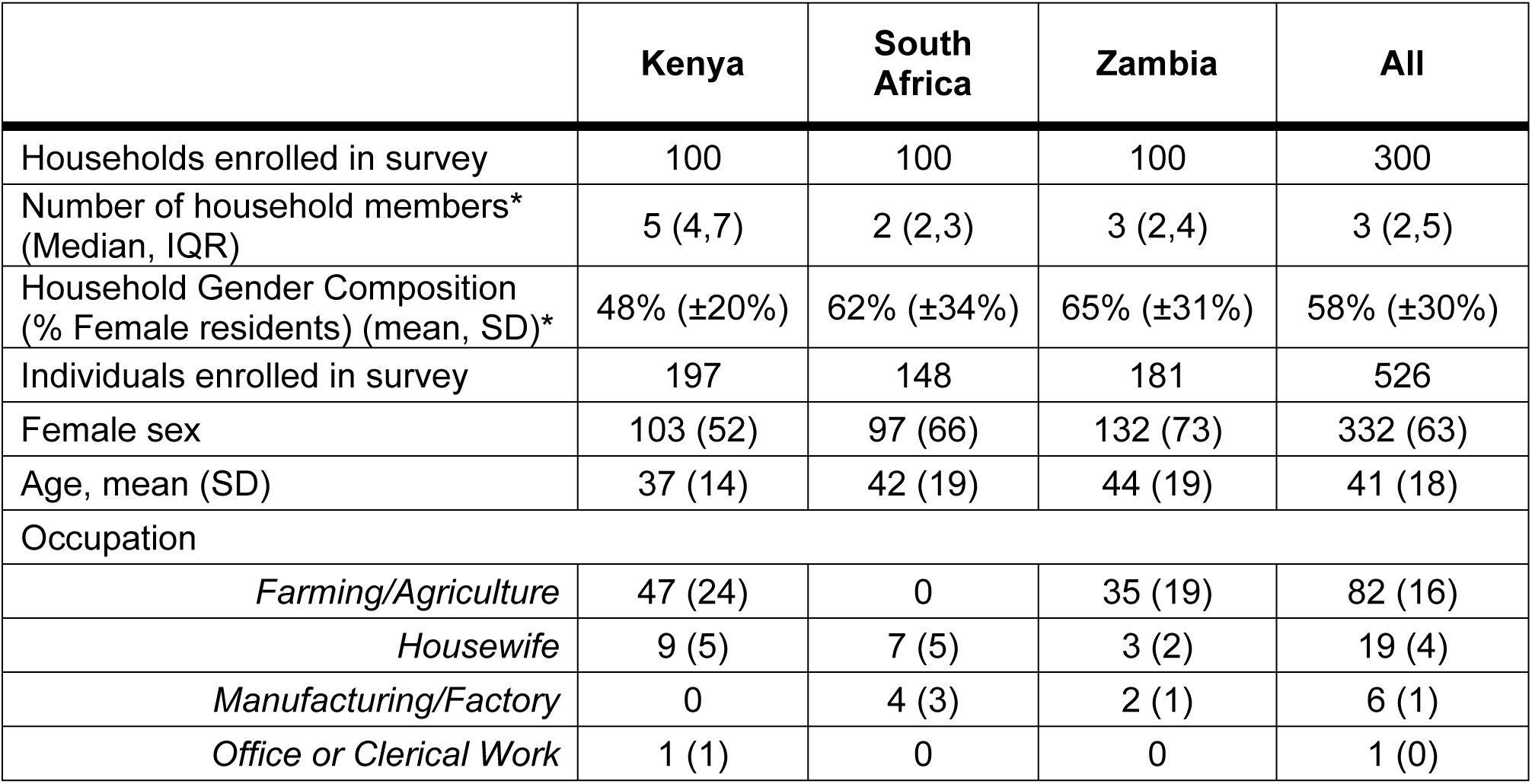
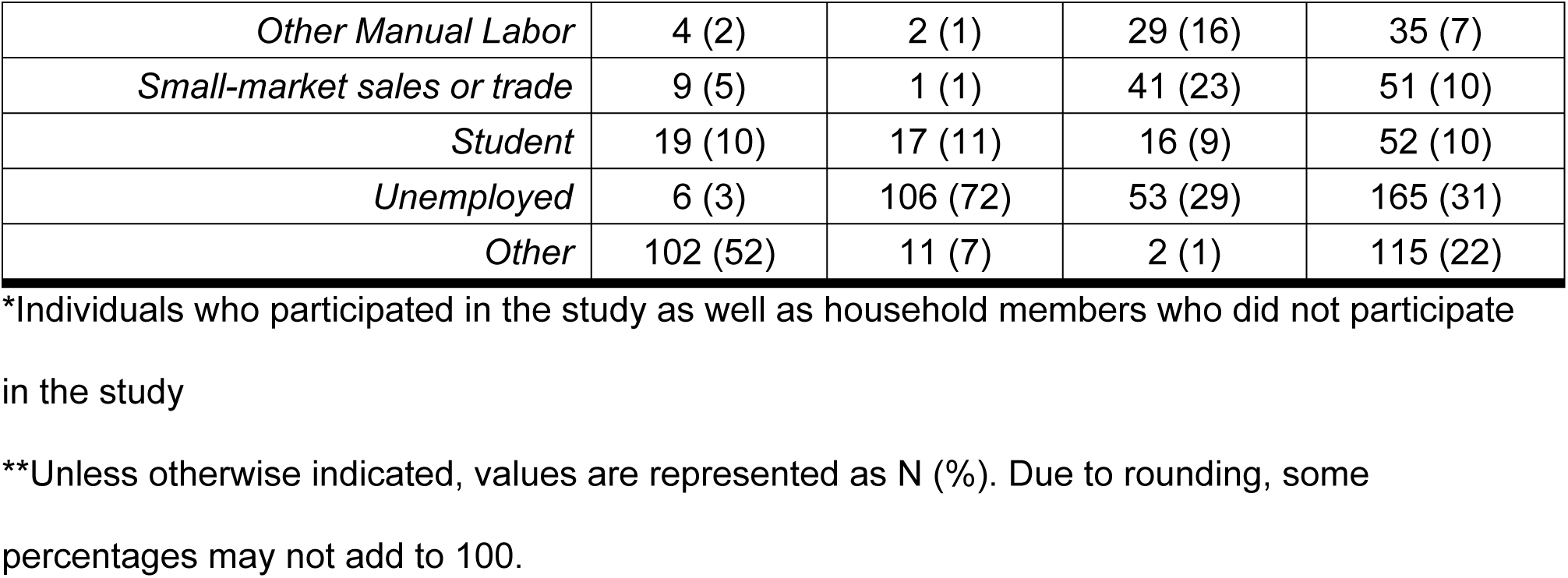
Demographics of Study Population.

### Feasibility of self-administered rapid diagnostic tests

Prior experience using a rapid test at home was low — 13% across all sites (Table 2). Despite limited experience with rapid tests, more than 90% of participants reported that the app-based instructions were “very easy” or “fairly easy” to understand and that conducting the test was “very easy” or “fairly easy” for HIV oral swab, HIV finger prick, and blood glucose finger prick tests (Fig 1a). All participants who self-tested for HIV, using either finger prick or oral swab tests, accurately interpreted their test results, with over 95% of participants reporting that they were “very confident” or “fairly confident” with their interpretation (Fig 1b).

**Fig 1.**
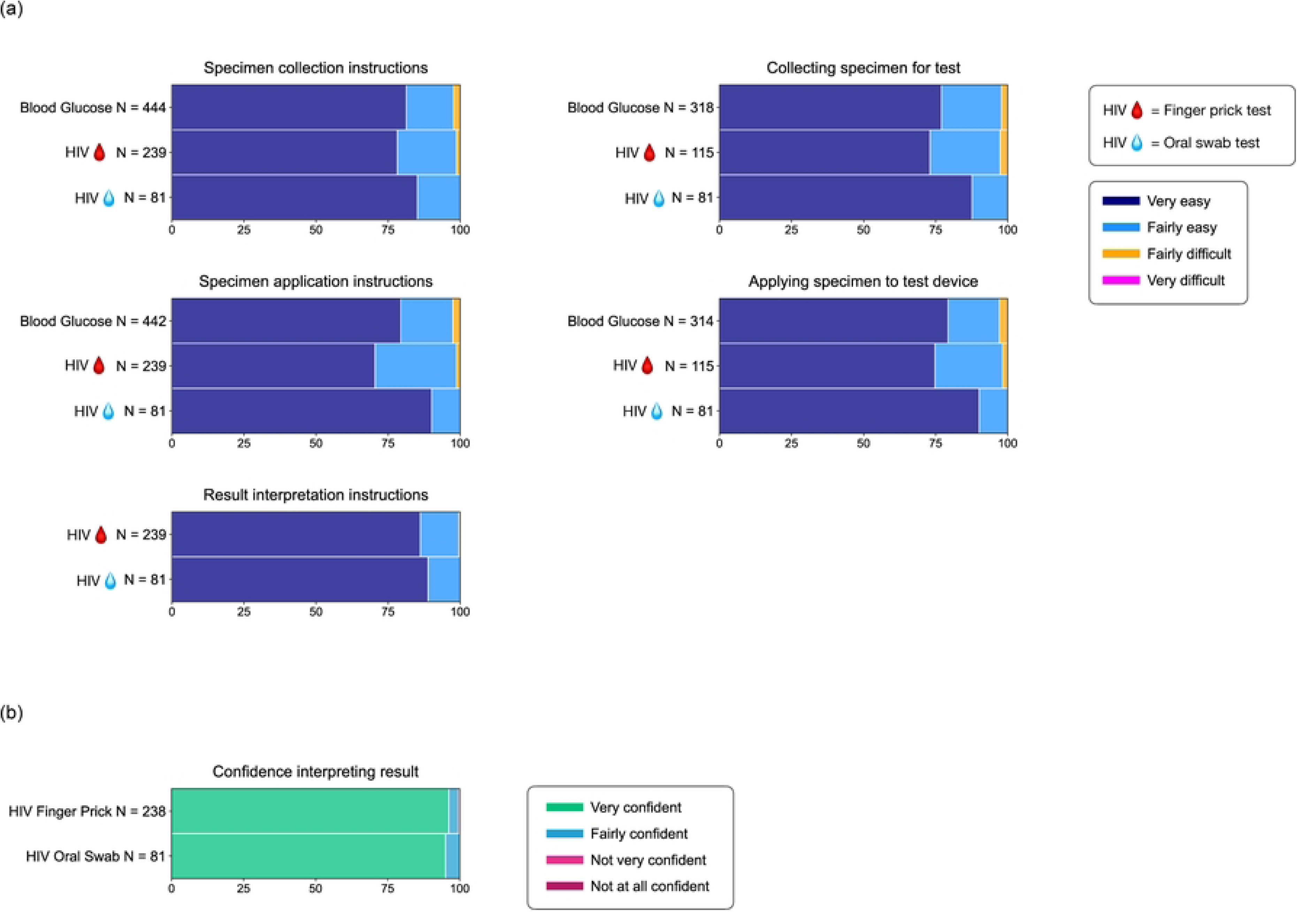
Feasibility of and preferences for different rapid test types. (a) Ease of understanding test instructions or completing test, (b) Confidence interpreting HIV test When asked about their preferences for diagnostic testing and disease monitoring, most participants preferred home-based testing (88%) versus clinic-based testing (9%) (Table 2).

**Table 2.** Acceptability of and preferences for any type of rapid test.

|  | **Kenya**<br>**N=197** | **South Africa**<br>**N=148** | **Zambia**<br>**N=181** | **All**<br>**N=526** |
| --- | --- | --- | --- | --- |
| Self-tested with a rapid test prior to the study* | 34 (17) | 32 (22) | 7 (4) | 73 (14) |
| Preference for where to self-test with a rapid test in the future <sup>†</sup> |  |  |  |  |
| *I would prefer to do this at home* | 168 (85) | 141 (95) | 149 (82) | 458 (87) |
| *I would prefer to do this in a clinical setting (e.g. at a local health clinic or hospital)* | 15 (8) | 1 (1) | 30 (17) | 46 (9) |
| *No preference/Anywhere* | 12 (6) | 1 (1) | 0 | 13 (2) |
| *Unsure/Don't know* | 0 | 0 | 2 (1) | 2 (<1) |
| Willing to use finger prick test on a 5-11 year old child? <sup>†</sup> |  |  |  |  |
| Yes | 147 (75) | 60 (41) | 131 (72) | 338 (64) |
| *Some children but not all children* | 21 (11) | 20 (14) | 9 (5) | 50 (10) |
| No | 22 (11) | 7 (5) | 37 (20) | 66 (13) |
| *Unsure / don't know* | 5 (3) | 56 (38) | 4 (2) | 65 (12) |
| Willing to use a finger prick test on a 12-15 year old child? <sup>†</sup> |  |  |  |  |
| Yes | 170 (86) | 75 (51) | 134 (74) | 379 (72) |
| *Some children but not all children* | 7 (4) | 14 (9) | 7 (4) | 28 (5) |
| No | 13 (7) | 2 (1) | 36 (20) | 51 (10) |
| *Unsure / don't know* | 5 (3) | 52 (35) | 4 (2) | 61 (12) |
| Willing to use this app or similar to take photos of test result and share with a community health worker in the future? | 192 (97) | 127 (86) | 177 (99) | 496 (95) |
| Willing to use this app or similar to take photos of test result and share with the Ministry of Health in the future? | 193 (98) | 116 (78) | 175 (98) | 484 (93) |

\* Unless otherwise indicated, values are represented as N (%). Due to rounding, some percentages may not add to 100.
<sup>†</sup> Responses were not obtained from 2 participants in Kenya and 5 participants in South Africa, thus total percentages for questions in these study sites will not add to 100.

Of participants who used an HIV oral swab test and a blood glucose finger prick test, 71% said they would be comfortable using either type of test (oral swab or finger prick) on their own in the future, whereas 23% reported they would only be comfortable using an oral swab test on their own in the future (Fig 2). For participants who used finger prick tests for both HIV and blood glucose tests, 37% reported they would be comfortable using either type of test (oral swab or finger prick) on their own in the future, with 53% of participants reporting they would only be comfortable with using a finger prick test in the future. Fewer than 5% of participants in the study population reported they would not be comfortable or were unsure if they would be comfortable conducting a finger prick or oral swab test on their own in the future.

**Fig 2.**
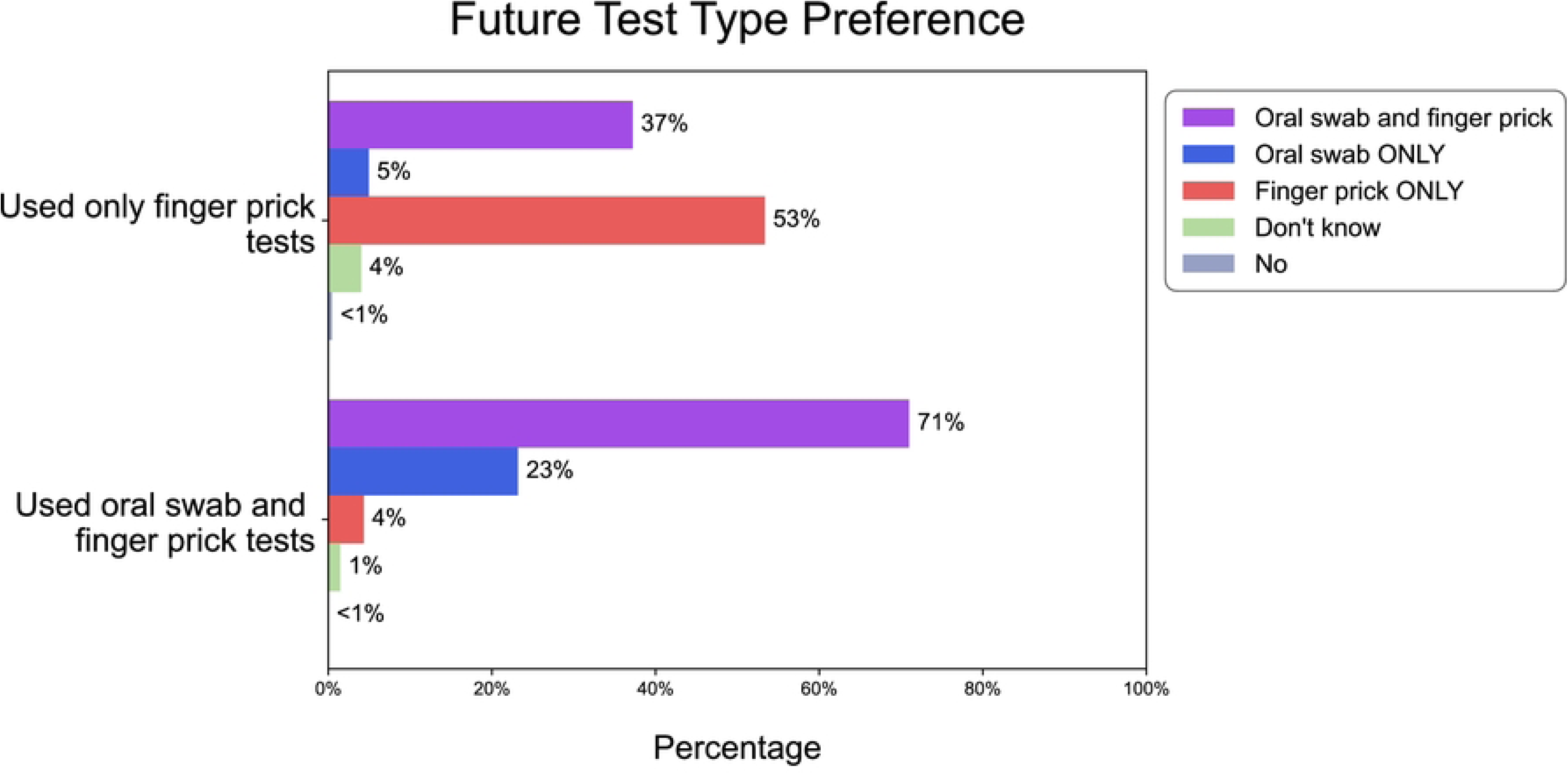
Preference for future self-test test type, based on test type used.

Participants also indicated a willingness to share self-test results in the future via a mobile app with community health workers (95%) and their respective Department/Ministry of Health (93%; Table 2).

### Prior HIV testing and HIV prevalence

The proportion of participants who were aware of living with HIV prior to study start varied across the study sites, with 14% in Kenya, 25% in South Africa, and 10% in Zambia (Table 3). Among participants who did not report a positive HIV status prior to study enrollment, 5% in Kenya, 4% in South Africa, and 15% in Zambia had never been tested for HIV and 39% in Kenya, 50% in South Africa, and 46% in Zambia had not been tested for HIV in over 12 months. For all participants without a known positive HIV status prior to the study, 68% in Kenya, 45% in South Africa, and 72% in Zambia consented to test for HIV and between 1-3% of those participants received a positive HIV rapid test and were referred for confirmatory testing (Table 4).

**Table 3.**
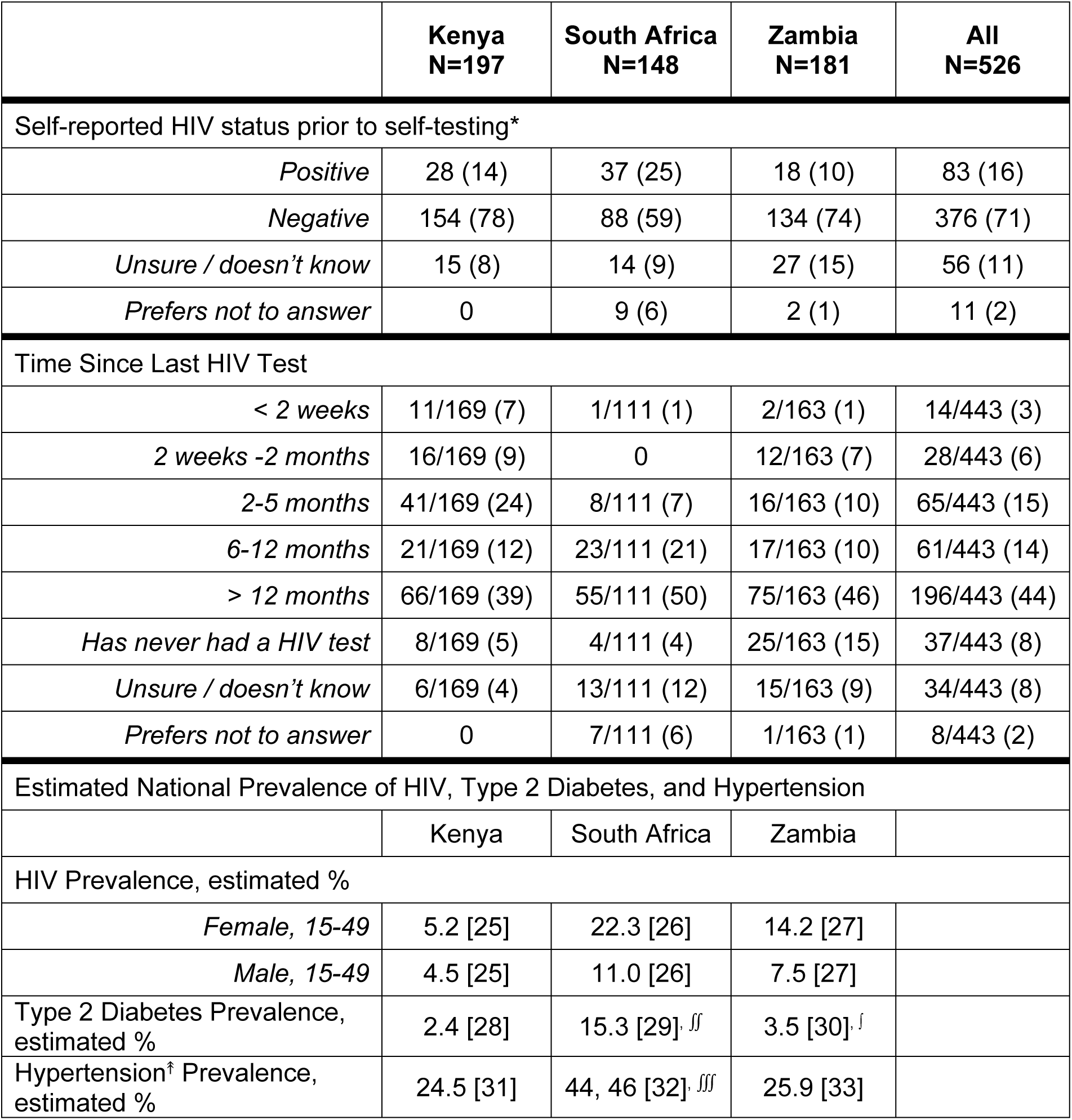
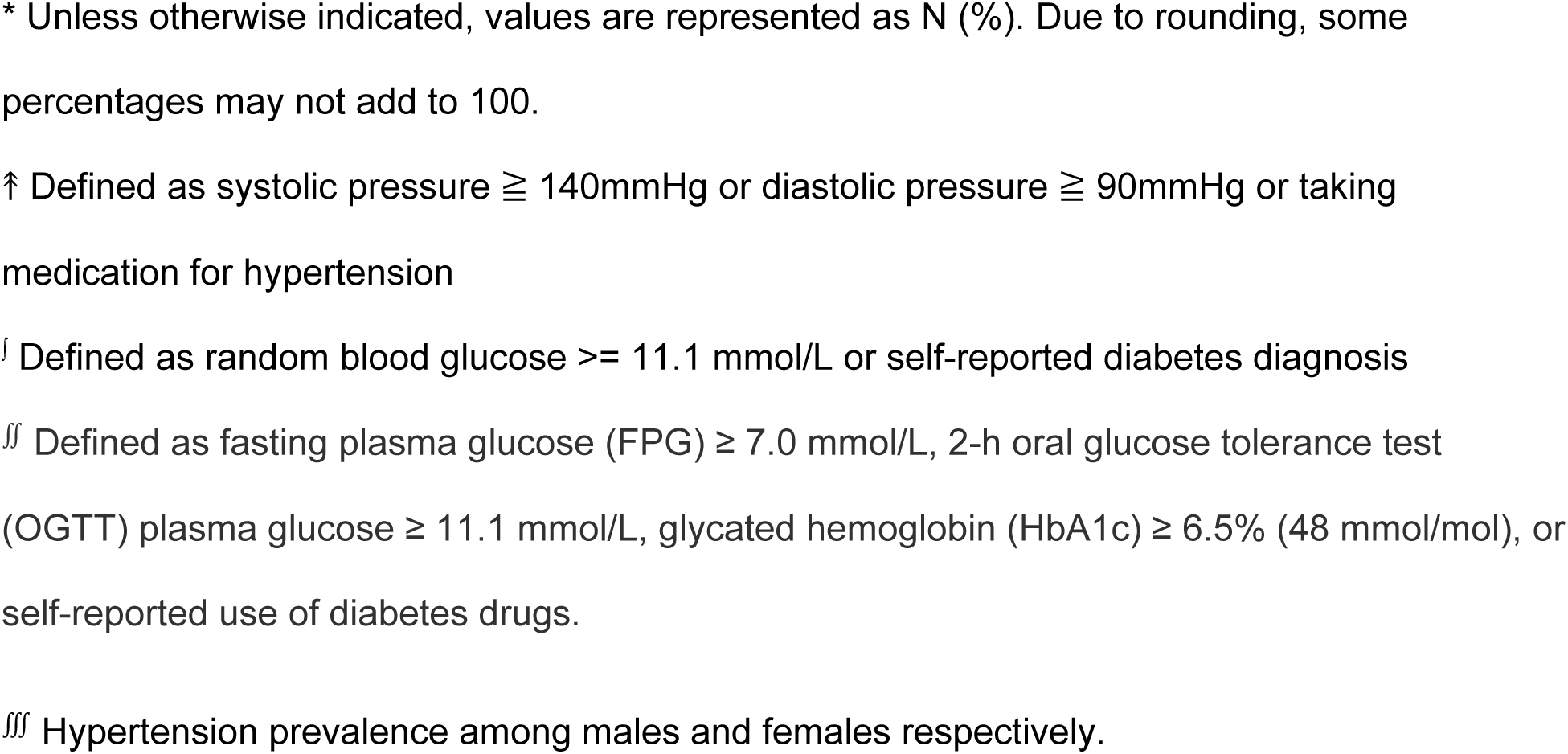
HIV Status and Testing History and Estimated National Prevalence of HIV, Type 2 Diabetes, and Hypertension.

|  | Kenya<br>N=197 | South Africa<br>N=148 | Zambia<br>N=181 | All<br>N=526 |
| --- | --- | --- | --- | --- |
| Self-reported HIV status prior to self-testing* |  |  |  |  |
| *Positive* | 28 (14) | 37 (25) | 18 (10) | 83 (16) |
| *Negative* | 154 (78) | 88 (59) | 134 (74) | 376 (71) |
| *Unsure / doesn't know* | 15 (8) | 14 (9) | 27 (15) | 56 (11) |
| *Prefers not to answer* | 0 | 9 (6) | 2 (1) | 11 (2) |
| Time Since Last HIV Test |  |  |  |  |
| *< 2 weeks* | 11/169 (7) | 1/111 (1) | 2/163 (1) | 14/443 (3) |
| *2 weeks -2 months* | 16/169 (9) | 0 | 12/163 (7) | 28/443 (6) |
| *2-5 months* | 41/169 (24) | 8/111 (7) | 16/163 (10) | 65/443 (15) |
| *6-12 months* | 21/169 (12) | 23/111 (21) | 17/163 (10) | 61/443 (14) |
| *> 12 months* | 66/169 (39) | 55/111 (50) | 75/163 (46) | 196/443 (44) |
| *Has never had a HIV test* | 8/169 (5) | 4/111 (4) | 25/163 (15) | 37/443 (8) |
| *Unsure / doesn't know* | 6/169 (4) | 13/111 (12) | 15/163 (9) | 34/443 (8) |
| *Prefers not to answer* | 0 | 7/111 (6) | 1/163 (1) | 8/443 (2) |
| Estimated National Prevalence of HIV, Type 2 Diabetes, and Hypertension |  |  |  |  |
|  | Kenya | South Africa | Zambia |  |
| HIV Prevalence, estimated % |  |  |  |  |
| *Female, 15-49* | 5.2 [25] | 22.3 [26] | 14.2 [27] |  |
| *Male, 15-49* | 4.5 [25] | 11.0 [26] | 7.5 [27] |  |
| Type 2 Diabetes Prevalence, estimated % | 2.4 [28] | 15.3 [29].<sup>ff</sup> | 3.5 [30].<sup>f</sup> |  |
| Hypertension<sup>†</sup> Prevalence, estimated % | 24.5 [31] | 44, 46 [32].<sup>fff</sup> | 25.9 [33] |  |

\* Unless otherwise indicated, values are represented as N (%). Due to rounding, some percentages may not add to 100.
† Defined as systolic pressure $\geq 140$ mmHg or diastolic pressure $\geq 90$ mmHg or taking medication for hypertension
‡ Defined as random blood glucose $\geq 11.1$ mmol/L or self-reported diabetes diagnosis
§ Defined as fasting plasma glucose (FPG) $\geq 7.0$ mmol/L, 2-h oral glucose tolerance test (OGTT) plasma glucose $\geq 11.1$ mmol/L, glycated hemoglobin (HbA1c) $\geq 6.5\%$ (48 mmol/mol), or self-reported use of diabetes drugs.
¶ Hypertension prevalence among males and females respectively.

**Table 4.** Results of rapid diagnostic tests for HIV, blood glucose and blood pressure.

|  | **Kenya**<br>**N=197** | **South Africa**<br>**N=148** | **Zambia**<br>**N=181** | **All**<br>**N=526** |
| --- | --- | --- | --- | --- |
| Tested for HIV* | 133 (68) | 66 (45) | 131 (72) | 330 (63) |
| *Negative* | 129/133 (97) | 65/66 (99) | 130/131 (99) | 324/330 (98) |
| *Positive* | 4/133 (3) | 1/66 (2) | 1/131 (1) | 6/330 (2) |
| Did not test for HIV because... | 64/197 (32) | 82/148 (55) | 50/181 (28) | 196/526 (37) |
| *participant previously tested positive for HIV<sup>‡</sup>* | 28/64 (44) | 24/82 (29) | 17/50 (34) | 69/196 (35) |
| *participant declined<sup>‡</sup>* | 28/64 (44) | 55/82 (67) | 21/50 (42) | 104/196 (53) |
| *test stockouts* | 0 | 1/82 (1) | 0 | 1/196 (1) |
| *other reason* | 8/64 (13) | 2/82 (2) | 12/50 (24) | 22/196 (11) |
| Tested blood glucose<sup>†</sup> | 177 (90) | 136 (92) | 156 (86) | 469 (89) |
| Blood glucose (mmol/L) (mean, SD) | 6.1 (2.6) | 6.6 (2.0) | 6.2 (1.5) | 6.3 (2.1) |
| Low blood glucose ($\leq 3.9$ mmol/L) | 3/177 (2) | 2/136 (2) | 2/156 (1) | 7/469 (1) |
| High blood glucose ($> 10$ mmol/L) | 5/177 (3) | 8/136 (6) | 5/156 (3) | 18/469 (4) |
| Did not test blood glucose because... | 20 (10) | 12 (8) | 25 (14) | 57 (11) |
| *participant declined* | 20/20 (100) | 10/12 (83) | 21/25 (84) | 51/57 (89) |
| *could not get enough blood from finger prick* | 0 | 0 | 4/25 (16) | 4/57 (7) |
| *research team ran out of test / materials* | 0 | 1/12 (8) | 0 | 1/57 (2) |
| *other reason* | 0 | 1/12 (8) | 0 | 1/57 (2) |
| Tested blood pressure | 196 (99) | 133 (90) | 173 (96) | 502 (95) |
| Systolic BP (mmHg) (mean, SD) | 118.8 (17) | 133.3 (20) | 134.0 (29) | 127.9 (24) |
| Diastolic BP (mmHg) (mean, SD) | 76.8 (10) | 79.6 (12) | 85.6 (15) | 80.6 (13) |
| High BP (systolic ≥ 140mmHg or diastolic ≥ 90mmHg) | 24/196 (12) | 38/133 (29) | 69/173 (40) | 131/502 (26) |

\* Unless otherwise indicated, values are represented as N (%). Due to rounding, some percentages may not add to 100.
‡ Participants who self-reported they were living with HIV either responded “previously tested positive for HIV” or “participant declined” and thus the total number of “previously tested positive for HIV” does not match the total participants who reported positive HIV status in Table 3.
† Participants self-tested for blood glucose during household visits that were conducted using convenience sampling, and therefore were not necessarily fasting before the blood glucose test.

### Blood glucose and blood pressure testing

Over 85% of participants at all study sites consented to blood glucose self-testing (Table 4). Between 3%-6% of participants had high blood glucose readings (> 10 mmol/L) and between 1%-2% of participants had low blood glucose readings (≤ 3.9 mmol/L).

Greater than 90% of participants across study sites consented to having their blood pressure measured by a member of the research team. Prevalence of high blood pressure varied across sites, with 12% in Kenya, 29% in South Africa and 40% in Zambia (Table 4).

## Discussion

In this cohort of adolescents and adults in peri-urban and rural areas of Kenya, South Africa, and Zambia, home-based, self-testing was both a feasible and preferred method to test for HIV, diabetes and hypertension. Despite few participants having experience with self-testing using a rapid test, most reported that home-based, self-testing was usable and acceptable for both finger prick blood and oral swab saliva specimen collection. Participants overwhelmingly reported a preference to conduct self-testing at home rather than self-testing in a clinic-setting. Participants who conducted self-tests using both oral swab and finger prick test types reported a greater comfort using either test type in the future. Participants who were given only finger prick tests were more likely to report comfort using only finger prick tests in the future. Gaining experience using oral swab tests seems to increase an individual’s comfort using that test type on their own in the future. This finding suggests that larger home-based, self-testing programs, which may struggle with rapid test availability, can implement programs by pooling multiple test types.

While our study was the first to evaluate feasibility of home-based self-testing for communicable diseases *and* NCDs, our findings are consistent with HIV home-based self-testing studies that have demonstrated a high degree of feasibility for rapid self-testing and a preference for home-based testing over clinic-based testing. [11,16,22,34,35] Previous studies offer mixed results for preference between HIV finger prick tests versus HIV oral swab tests, when given the option. [36,37] Our study found that when participants were given the opportunity to use both test types, 69% were comfortable using either test type in the future. Additionally, we confirmed that individuals find blood glucose self-testing is highly feasible, which is consistent with other studies evaluating self-monitored blood glucose (SMBG) programs in LMICs. [38,39]

Our study sites represent the regional variability in HIV, high blood glucose, and high blood pressure prevalence within each of their respective countries. We found comparable post-study prevalence of HIV in study participants at the South Africa (26%) and Zambia (10%) sites when compared to national estimates. In Kenya, 16% of participants either had a positive HIV test or had previously screened positive for HIV, higher than national estimates of 5.2% for females and 4.5% for males but consistent with prevalence in the sampled Migori County (Table 3). We found a comparable prevalence of high blood glucose among study participants in Kenya (3%) and Zambia (3%) compared to national estimates for diabetes. In South Africa, 6% of participants’ tests met the threshold for high blood glucose whereas national estimates of diabetes are much higher at 15.3% (Table 3). The percentage of participants with high blood pressure readings was 12%, 29% and 40% in Kenya, South Africa and Zambia respectively. The national estimates for hypertension in Kenya, South Africa, and Zambia are 25%, 45.0%, and 25.9% respectively (Table 3). Our findings and the national estimates together indicate that HIV, high blood sugar, and high blood pressure are major health issues in each of these countries.

Interpreting continuous values like blood glucose and blood pressure, which can fluctuate throughout the day, will require greater patient education, especially given that diabetes and hypertension are both new public health issues and are widely undiagnosed and untreated in the study sites. [40,41] Isolated studies of patient education and disease management programs for diabetes and hypertension self-monitoring in LMICs have been successful. [38,39,42,43]

Addressing the availability and affordability of rapid tests approved for self-use is crucial for the sustainability of self-testing programs. One of the strengths of our study was eliminating typical barriers to access to rapid tests by providing the rapid test and study mobile devices with a digital tool so that we could study feasibility of the tests themselves.

The willingness of participants to share their results with community health workers and Ministries/Departments of Health signals an opportunity for future research to explore the design and implementation of self-testing public health surveillance programs in LMICs.

Our study had several strengths and limitations. Unlike many other feasibility studies which have evaluated rapid tests for a single, communicable disease like HIV, this study evaluated tests for communicable and NCDs in the same study population. This study also demonstrated the feasibility of multiple types of rapid tests, including finger prick blood and oral swab saliva tests, across multiple study sites in sub-Saharan Africa, indicating that future research and disease monitoring programs can likely rely on pooling test types for efficient and user-friendly self-testing.

Due to its cross-sectional nature, this study’s single timepoint blood pressure testing was not meant to be diagnostic for hypertension. Likewise, the use of blood glucose tests without a fasting protocol was not diagnostic of diabetes. Additionally, while many participants reported comfort sharing future self-test results with either community health workers or a Ministry/Department of Health, social desirability bias may have influenced their survey response. Future research allowing participants the opportunity to share their results with relevant health authorities can better estimate this willingness to share results for public health efforts.

## Conclusion

Home-based rapid testing can empower individuals to self-test for communicable and NCDs. The results of this study indicate that self-testing for HIV and blood glucose is a feasible and preferred method for individuals in the sites studied. Self-testing alerts individuals as to whether or not they need to seek confirmatory testing in a clinic-based setting and, potentially, future treatment. Our findings both provide evidence that individuals value home-based self-testing, and support efforts to broaden the application of self-testing to more diseases in LMICs.

## Data Availability

The de-identified data underlying the findings of this study are available in the Harvard Dataverse repository at DOI: https://doi.org/10.7910/DVN/BKU6VK. Access to the dataset is restricted due to the sensitive nature of the information collected, including participant HIV status, and to protect participant anonymity. Researchers who meet the criteria for access to confidential data may request access by contacting the study team via the repository link.

## Acknowledgements

The authors thank Dr. Sasha Frade for her insightful review of the manuscript, Paul Isabelli and Sarah Morris for their contributions to the study’s conception and support throughout, and the developers at Audere for creating the HealthPulse TestNow application used in this study. We also thank the study participants who contributed to this research.

## Notes

### Competing Interest Statement

The authors have declared no competing interest.

### Funding Statement

This work was supported by the Gates Foundation, grant number INV-063459. The conclusions and opinions expressed in this work are those of the author(s) alone and shall not be attributed to the Foundation.

### Author Declarations

The study was reviewed and approved by the University of Washington Institutional Review Board, Human Subjects Division; the Human Sciences Research Council (HSRC) Research Ethics Committee in South Africa, the Scientific and Ethics Review Unit at the Kenya Medical Research Institute, and ERES Converge IRB in Zambia.

## REFERENCES

[1] Boutayeb A. The Burden of Communicable and Non-Communicable Diseases in Developing Countries BT - Handbook of Disease Burdens and Quality of Life Measures. Handbook of Disease Burdens and Quality of LIfe Measures, 2010.

[2] Institute for Health Metrics and Evaluation. Global Burden of Disease Results Tool. 2019. http://ghdx.healthdata.org/gbd-results-tool (accessed March 20, 2024).

[3] Jayatilleke K. Challenges in Implementing Surveillance Tools of High-Income Countries (HICs) in Low Middle Income Countries (LMICs). Curr Treat Options Infect Dis 2020;12. 10.1007/s40506-020-00229-2.

[4] Srivastava D, McGuire A. Patient access to health care and medicines across low-income countries. Soc Sci Med 2015;133. 10.1016/j.socscimed.2015.03.021.

[5] McElfish PA, Purvis R, James LP, Willis DE, Andersen JA. Perceived barriers to covid-19 testing. Int J Environ Res Public Health 2021;18. 10.3390/ijerph18052278.

[6] Gilbert M, Thomson K, Salway T, Haag D, Grennan T, Fairley CK, et al. Differences in experiences of barriers to STI testing between clients of the internet-based diagnostic testing service GetCheckedOnline.com and an STI clinic in Vancouver, Canada. Sex Transm Infect 2019;95. 10.1136/sextrans-2017-053325.

[7] Fleming E, Oremo J, O’Connor K, Odhiambo A, Ye T, Oswago S, et al. The Impact of Integration of Rapid Syphilis Testing during Routine Antenatal Services in Rural Kenya. J Sex Transm Dis 2013;2013. 10.1155/2013/674584.

[8] Wynn A, Ramogola-Masire D, Gaolebale P, Moshashane N, Agatha Offorjebe O, Arena K, et al. Acceptability and Feasibility of Sexually Transmitted Infection Testing and Treatment among Pregnant Women in Gaborone, Botswana, 2015. Biomed Res Int 2016;2016. 10.1155/2016/1251238.

[9] Martin K, Wenlock R, Roper T, Butler C, Vera JH. Facilitators and barriers to point-of-care testing for sexually transmitted infections in low- and middle-income countries: a scoping review. BMC Infect Dis 2022;22. 10.1186/s12879-022-07534-9.

[10] Choko AT, MacPherson P, Webb EL, Willey BA, Feasy H, Sambakunsi R, et al. Uptake, Accuracy, Safety, and Linkage into Care over Two Years of Promoting Annual Self-Testing for HIV in Blantyre, Malawi: A Community-Based Prospective Study. PLoS Med 2015;12:e1001873. 10.1371/JOURNAL.PMED.1001873.

[11] Majam M, Mazzola L, Rhagnath N, Lalla-Edward ST, Mahomed R, Venter WDF, et al. Usability assessment of seven HIV self-test devices conducted with lay-users in Johannesburg, South Africa. PLoS One 2020;15. 10.1371/journal.pone.0227198.

[12] Reid SD, Fidler SJ, Cooke GS. Tracking the progress of HIV: The impact of point-of-care tests on antiretroviral therapy. Clin Epidemiol 2013;5. 10.2147/CLEP.S37069.

[13] World Health Organization. Implementation of Self-Care Interventions for Health and Well-Being: Guidance for Health Systems. ISBN 9789240094888. Geneva: 2024.

[14] Vrana-Diaz CJ, Korte JE, Gebregziabher M, Richey L, Selassie A, Sweat M, et al. Relationship Gender Equality and Couples’ Uptake of Oral Human Immunodeficiency Virus Self-Testing Kits Delivered by Pregnant Women in Kenya. Sex Transm Dis 2019;46. 10.1097/OLQ.0000000000001037.

[15] Adjiwanou V, LeGrand T. Gender inequality and the use of maternal healthcare services in rural sub-Saharan Africa. Health Place 2014;29. 10.1016/j.healthplace.2014.06.001.

[16] Figueroa C, Johnson C, Ford N, Sands A, Dalal S, Meurant R, et al. Reliability of HIV rapid diagnostic tests for self-testing compared with testing by health-care workers: a systematic review and meta-analysis. Lancet HIV 2018;5. 10.1016/S2352-3018(18)30044-4.

[17] Lindner AK, Nikolai O, Rohardt C, Kausch F, Wintel M, Gertler M, et al. Diagnostic accuracy and feasibility of patient self-testing with a SARS-CoV-2 antigen-detecting rapid test. Journal of Clinical Virology 2021;141. 10.1016/j.jcv.2021.104874.

[18] Jones HE, Altini L, De Kock A, Young T, Van De Wijgert JHHM. Home-based versus clinic-based self-sampling and testing for sexually transmitted infections in Gugulethu, South Africa: Randomised controlled trial. Sex Transm Infect 2007;83. 10.1136/sti.2007.027060.

[19] Elmardi KA, Malik EM, Abdelgadir T, Ali SH, Elsyed AH, Mudather MA, et al. Feasibility and acceptability of home-based management of malaria strategy adapted to Sudan’s conditions using artemisinin-based combination therapy and rapid diagnostic test. Malar J 2009;8. 10.1186/1475-2875-8-39.

[20] Mukoka M, Sibanda E, Watadzaushe C, Kumwenda M, Abok F, Corbett EL, et al. COVID-19 self-testing using antigen rapid diagnostic tests: Feasibility evaluation among health-care workers and general population in Malawi. PLoS One 2023;18. 10.1371/journal.pone.0289291.

[21] Hergott DEB, Owalla TJ, Balkus JE, Apio B, Lema J, Cemeri B, et al. Feasibility of community at-home dried blood spot collection combined with pooled reverse transcription PCR as a viable and convenient method for malaria epidemiology studies. Malar J 2022;21. 10.1186/s12936-022-04239-x.

[22] Sabapathy K, van den Bergh R, Fidler S, Hayes R, Ford N. Uptake of Home-Based Voluntary HIV Testing in Sub-Saharan Africa: A Systematic Review and Meta-Analysis. PLoS Med 2012;9. 10.1371/journal.pmed.1001351.

[23] Medina-Marino A, de Vos L, Bezuidenhout D, Denkinger CM, Schumacher SG, Shin SS, et al. “I got tested at home, the help came to me”: acceptability and feasibility of home-based TB testing of household contacts using portable molecular diagnostics in South Africa. Tropical Medicine and International Health 2021;26. 10.1111/tmi.13533.

[24] Kish L. A Procedure for Objective Respondent Selection within the Household. J Am Stat Assoc 1949;44. 10.1080/01621459.1949.10483314.

[25] National AIDS and STI Control Programme. Kenya HIV Estimates Report 2018. 2018.

[26] Human Sciences Research Council (HSRC) C for DC. Sixth South African National HIV Prevalence, Incidence, and Behaviour survey. Pretoria: 2023.

[27] Zambia Statistics Agency M of HZ and ICF. Zambia Demographic and Health Survey 2018. Lusaka, Zambia: 2019.

[28] Mohamed SF, Mwangi M, Mutua MK, Kibachio J, Hussein A, Ndegwa Z, et al. Prevalence and factors associated with pre-diabetes and diabetes mellitus in Kenya: Results from a national survey. BMC Public Health 2018;18. 10.1186/s12889-018-6053-x.

[29] Pheiffer C, Wyk VP Van, Turawa E, Levitt N, Kengne AP, Bradshaw D. Prevalence of type 2 diabetes in South Africa: A systematic review and meta-analysis. Int J Environ Res Public Health 2021;18. 10.3390/ijerph18115868.

[30] Bailey S Lou, Ayles H, Beyers N, Godfrey-Faussett P, Muyoyeta M, du Toit E, et al. Diabetes mellitus in Zambia and the Western Cape province of South Africa: Prevalence, risk factors, diagnosis and management. Diabetes Res Clin Pract 2016;118. 10.1016/j.diabres.2016.05.001.

[31] Mohamed SF, Mutua MK, Wamai R, Wekesah F, Haregu T, Juma P, et al. Prevalence, awareness, treatment and control of hypertension and their determinants: Results from a national survey in Kenya. BMC Public Health 2018;18. 10.1186/s12889-018-6052-y.

[32] Health ND of, F IC. South Africa Demographic and Health Survey 2016. Pretoria: National Department of Health - NDoH - ICF; 2019.

[33] Goma FM, Mwewa B, Tembo GK, Kachamba M, Syatalimi C, Simweemba C, et al. May Measurement Month 2017: Blood pressure screening results from Zambia - Sub-Saharan Africa. European Heart Journal, Supplement 2019;21. 10.1093/eurheartj/suz077.

[34] Choko AT, Desmond N, Webb EL, Chavula K, Napierala-Mavedzenge S, Gaydos CA, et al. The uptake and accuracy of oral kits for HIV self-testing in high HIV prevalence setting: A cross-sectional feasibility study in Blantyre, Malawi. PLoS Med 2011;8. 10.1371/journal.pmed.1001102.

[35] Pettifor A, Lippman SA, Kimaru L, Haber N, Mayakayaka Z, Selin A, et al. HIV self-testing among young women in rural South Africa: A randomized controlled trial comparing clinic-based HIV testing to the choice of either clinic testing or HIV self-testing with secondary distribution to peers and partners. EClinicalMedicine 2020;21. 10.1016/j.eclinm.2020.100327.

[36] Lippman SA, Gilmore HJ, Lane T, Radebe O, Chen YH, Mlotshwa N, et al. Ability to use oral fluid and fingerstick HIV self-testing (HIVST) among South African MSM. PLoS One 2018;13. 10.1371/journal.pone.0206849.

[37] Njau B, Covin C, Lisasi E, Damian D, Mushi D, Boulle A, et al. A systematic review of qualitative evidence on factors enabling and deterring uptake of HIV self-testing in Africa. BMC Public Health 2019;19. 10.1186/s12889-019-7685-1.

[38] Ng’ang’a L, Ngoga G, Dusabeyezu S, Hedt-Gauthier BL, Harerimana E, Niyonsenga SP, et al. Feasibility and effectiveness of self-monitoring of blood glucose among insulin-dependent patients with type 2 diabetes: open randomized control trial in three rural districts in Rwanda. BMC Endocr Disord 2022;22. 10.1186/s12902-022-01162-9.

[39] Pastakia SD, Cheng SY, Kirui NK, Kamano JH. Dynamics, impact, and feasibility of self-monitoring of blood glucose in the rural, resource-constrained setting of western Kenya. Clinical Diabetes 2015;33. 10.2337/diaclin.33.3.136.

[40] Manne-Goehler J, Atun R, Stokes A, Goehler A, Houinato D, Houehanou C, et al. Diabetes diagnosis and care in sub-Saharan Africa: pooled analysis of individual data from 12 countries. Lancet Diabetes Endocrinol 2016;4. 10.1016/S2213-8587(16)30181-4.

[41] Mbanya JCN, Motala AA, Sobngwi E, Assah FK, Enoru ST. Diabetes in sub-Saharan Africa. The Lancet 2010;375. 10.1016/S0140-6736(10)60550-8.

[42] Lamptey R, Robben MP, Amoakoh-Coleman M, Boateng D, Grobbee DE, Davies MJ, et al. Structured diabetes self-management education and glycaemic control in low- and middle-income countries: A systematic review. Diabetic Medicine 2022;39. 10.1111/dme.14812.

[43] Gill G V., Price C, Shandu D, Dedicoat M, Wilkinson D. An effective system of nurse-led diabetes care in rural Africa. Diabetic Medicine 2008;25. 10.1111/j.1464-5491.2008.02421.x.

